# Hospital air sampling enables surveillance of respiratory virus infections and genomes

**DOI:** 10.1101/2025.03.18.25324190

**Authors:** Sofiya Bobrovska, Erin Newcomer, Michael Gottlieb, V. Eloesa McSorley, Alyse Kittner, Mary K. Hayden, Stefan Green, Hannah J. Barbian

## Abstract

There is an urgent need for early detection and comprehensive surveillance of respiratory pathogens. Environmental surveillance may be key to timely responses for newly emerging pathogens and infections that are unreported or underreported. Here, we employed air sampling in a large urban hospital. Air samples (n=358) were collected weekly at five locations, including two in the emergency department, two in hospital common areas and one in a storage room, for two respiratory virus seasons (November 2022 to June 2024). Air samples were tested for eight respiratory pathogens by qPCR, including RNA and DNA viruses and a bacterium. Air samples had an average of four detected pathogens per sample and 97% samples contained SARS-CoV-2. Air sample pathogen positivity and quantity were strongly correlated with clinical surveillance for four seasonal respiratory pathogens: influenza A and B, respiratory syncytial virus, and human metapneumovirus. Targeted sequencing of SARS-CoV-2 showed that lineages detected in air samples reflected those in contemporaneous regional clinical specimens. Metagenomic sequencing with viral enrichment detected myriad human pathogens, including respiratory-associated viruses with recovery of full viral genomes. Detection of viral pathogens correlated well between metagenomic sequencing and qPCR. Overall, this suggests air sampling can be an agile and effective tool for pathogen early warning, surveillance and genome characterization.

## 1. Introduction

The recent SARS-CoV-2 pandemic and mpox outbreaks have highlighted the importance of early detection, ongoing surveillance and genome characterization of viruses to inform timely public health responses. Further, seasonal respiratory viruses resulting in significant annual morbidity, mortality and economic costs and have shown less predictable seasonality following the COVID-19 pandemic, underscoring the need for more comprehensive viral surveillance [1–4]. Accurate estimates of respiratory pathogen levels are critically important, especially in healthcare and other congregate settings that are at risk for outbreaks. However, clinical surveillance systems at both population- and building-scale can be difficult to organize, labor-intensive, and expensive. Further, detection by clinical testing can be insufficient when pathogens are newly emerging, when cases are largely unreported, or when unexpected or off-season virus outbreaks occur.

Environmental sampling does not rely on clinical testing and can provide an early warning for pathogen presence as well as estimation of community infection rates [5]. Wastewater sampling has been embraced for population-level SARS-CoV-2 surveillance, especially as clinical testing data declined following increased use of at-home antigen tests, “pandemic fatigue”, and changing case reporting requirements [6]. Wastewater sampling has also proved valuable for tracking SARS-CoV-2 variant dynamics [7]. However, wastewater-based surveillance is mostly limited to large population-scale analyses. Finer-scale neighborhood or building-level sewage can be difficult or impossible to access and rural sites with septic systems may not be able to be incorporated into wastewater surveillance programs.

Indoor air samples are easier to access environmental specimens and can provide pathogen surveillance at a building- or room-scale. Others have found that air sampling can be used to detect various bacterial and viral pathogens, but it has historically faced challenges such as low pathogen recovery due to limited biomass, tedious processing, and contamination. However, new air samplers have recently been developed that are more compact, inexpensive, and have simplified processing and efficient virus recovery, increasing ease of placement and accessibility to non-expert users. These have proven effective at detecting SARS-CoV-2 in controlled settings such as patient isolation rooms [8]. A few promising studies identified SARS-CoV-2 and other respiratory pathogens in community settings like coffee shops and schools using air sampling [9, 10].

Acute-care hospitals and emergency departments (EDs) represent high pathogen burden settings, where individuals with respiratory and infectious diseases are likely to seek care, and thus may serve as ideal locations for pathogen detection. Further, EDs include a population traditionally underrepresented in primary care settings and are a common location for those with novel or newly emerging pathogens to present [11–13]. Others have found that air samples from acute care hospitals and EDs contain respiratory viruses [14–16], however, it is not yet known whether air sample positivity can be extrapolated to regional clinical positivity.

Viral genome characterization can enrich surveillance data by allowing for identification of variants/subtypes, evolution and mutations of interest, and prediction of immune sensitivity, virulence, and transmissibility [17–19]. Further, genome sequences can help inform diagnostic, vaccine and therapeutic design [20–22]. Few studies have successfully characterized viral genomes from air samples, likely due to technical challenges associated with low biomass air samples [23]. SARS-CoV-2 directed sequencing studies have been applied in controlled settings like individual COVID inpatient rooms, [24] or targeting small genomic regions [9]. Existing studies performing metagenomic sequencing of indoor air viruses have often found low proportions of human viruses (<0.001%) in sequence data [25–28]. One study found promising results using single-strand, RNA-targeted amplification to recover several complete viral genomes from air samples [29]. Metagenomic capture enrichment techniques may improve viral recovery from environmental samples, but have thus far only been tested using wastewater environmental samples [30–32].

To address this gap, this study aimed to assess air sampling for pathogen detection and genomic surveillance, as a complementary or alternative surveillance tool when clinical and wastewater testing may be limited or challenging to deploy.

## 2. Materials and methods

### 2.1 Air sample collection and processing

This was a prospective, observational study using ambient air sampling for virus detection over a 19-month period (Fig 1). This study was found to not meet the definition of human subjects research and was deemed exempt by the Rush University Medical Center Institutional Review Board. AerosolSense samplers (Thermo Fisher Scientific) were placed at table-height (0.76 meters from the floor) at five locations within Rush University Medical Center (RUMC) in Chicago, IL [33]. RUMC is a 671 bed, urban quaternary care center. The ED at RUMC has a 70,000 patient per year annual volume with a 3-year emergency medicine residency program. Two air samplers were placed in the ED, with one ∼6 meters from the ED reception desk in the patient waiting room area and another ∼2 meters from a nurse’s charting station in an ED hallway used frequently for patient transport and temporary patient triage/overflow. Two were placed outside of the ED in hospital congregate areas, with one in a central hospital café seating area <1 meter from the nearest seats and one ∼3 meters from a visitor registration desk at a main hospital entrance. The fifth was placed in an infrequently accessed storage room as a negative control. Air samples were collected between Nov 10, 2022 and Jun 13, 2024, spanning two respiratory virus seasons defined as October 1 – March 31. AerosolSense cartridges (Thermo Fisher Scientific), which collect particles 0.1 – 15 μm in diameter, were inserted according to manufacturer instructions and air was sampled continuously for 7 days at the instrument default of 200 L/min. Previous studies show that 7-day samples was representative of positivity in daily samples taken over the same timeframe [9]. At the conclusion of the week of sampling, the cartridge was removed, transferred to the laboratory and processed within 24 hours of collection. Cartridge processing and extraction were adapted from those previously described [9]. Processing took place in a biosafety cabinet used exclusively for low-biomass pre-PCR specimens and thoroughly cleaned using UV light, 70% ethanol and RNase AWAY (Applied Biological Materials). The two membranes contained within each cartridge were removed using a disposable forceps and transferred to separate nucleic acid-free tubes containing 500 µl DNA/RNA Shield Stabilization Solution (Zymo Research). One membrane was then transferred to −80°C storage and one was further processed. Membranes were manually depressed using a pipette tip until dust particulates appeared fully suspended in the solution. 300 µl solution was then used for total nucleic acid extraction using the Maxwell RSC Viral Total Nucleic Acid Purification Kit and Maxwell RSC automated extraction instrument (Promega) according to manufacturer instructions. An extraction blank was included in every extraction run by processing an AerosolSense cartridge that had not been subjected to sampling. Nucleic acid extracts were eluted in 75 µl nuclease-free water, tested via qPCR within 24 hours and then stored at −80°C.

**Fig 1.**
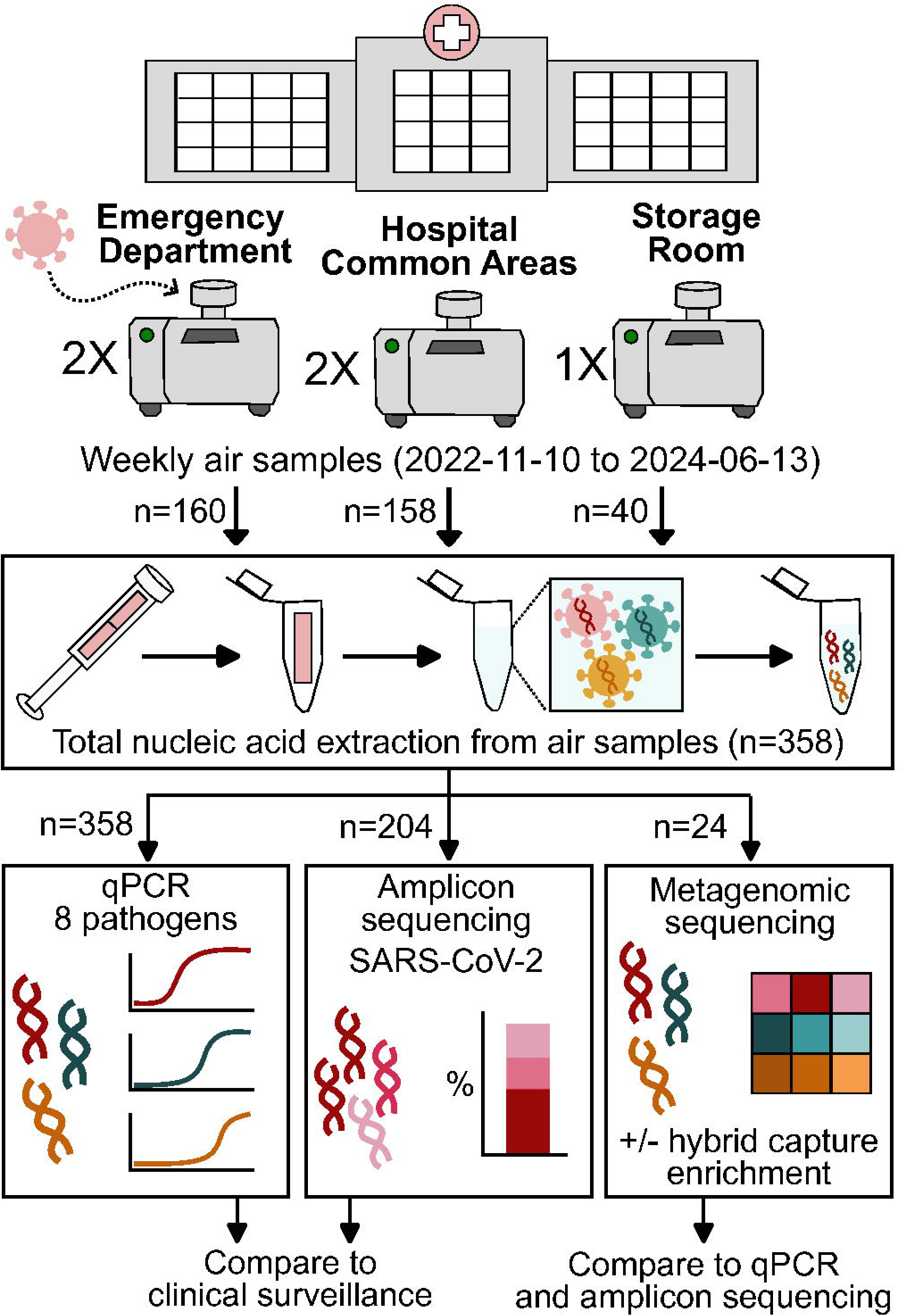
Air sample collection and analysis outline. Five air samplers were placed throughout a large urban acute-care hospital: two within the emergency department (one near reception and one in a hallway used for patient transport/triage), two within hospital common areas (one in a central café seating area and one near a visitor registration desk) and one within a storage room with minimum human activity. Weekly hospital air samples were collected and subjected to total nucleic acid extraction. All air sample nucleic acid extracts were analyzed by quantitative PCR (qPCR) for eight respiratory pathogens to assess pathogen presence and quantity: SARS-CoV-2, respiratory syncytial virus A/B, influenza A, influenza B, human metapneumovirus, adenovirus, enterovirus, and *Streptococcus pneumoniae*. Respiratory pathogen air detections and quantities were compared to clinical surveillance data at the national and hospital level. A subset (57%) of air samples were subjected to SARS-CoV-2 amplicon sequencing and lineage proportions were compared to clinical specimen sequence data. A small subset (7%) of air samples were subjected to metagenomic sequencing with and without hybrid capture enrichment of human pathogenic viruses. Metagenomic sequencing data was compared to pathogen detection, quantity, and lineage data obtained using qPCR and amplicon sequencing.

### 2.2 Respiratory pathogen qPCR detection

Total nucleic acid extracts were tested for the presence of selected viral and bacterial pathogens using a TrueMark custom plate in 96-well format (Thermo Fisher Scientific). Pathogen-specific assay IDs selected were Vi99990014_po/Vi07922149_s1 (RSV A/B), Vi07921935_s1 (SARS-CoV-2 ORF1ab), Vi99990011_po (pan influenza A), Vi07922143_s1/Vi07922144_s1 (influenza B), Vi99990004_po (human metapneumovirus), Vi07922136_s1/ Vi07922137_s1 (adenovirus), Vi06439631_s1 (pan enterovirus) and Ba06439619_s1 (*Streptococcus penumoniae*). All wells also contained a positive control assay for human RNaseP (assay ID RNAP_CDC_QSY_JUN). TrueMark plates were centrifuged at 2000xg for 1 min then 5 µl TaqPath 1-Step Multiplex Master Mix (Thermo Fisher Scientific) diluted to manufacturer instructions and 5 µl air sample extract were added to each well. Plates were sealed, vortexed, centrifuged at 1400xg for 20 sec, then run on a Quantstudio 5 real-time PCR instrument with standard cycling mode (Thermo Fisher Scientific). Cycling conditions were 25°C for 2 minutes, 53°C for 10 min, 85°C for 10 min, 95°C for 2 min, then 40 cycles of 95°C for 3 sec, 60°C for 30 sec, followed by a final step of 60°C for 30 sec. Constant baseline and threshold values, determined through limit of detection studies below, were applied and data were exported from the Quantstudio Design & Analysis Software v.2.6.0 and imported into RStudio for further analysis [34]. The following quality controls were used: i) an extraction blank was processed with every qPCR plate, extractions were repeated or excluded if amplification above thresholds was detected in the extraction blank, ii) within-sample RNaseP Ct values were compared as internal control, samples with standard deviation above 1.5 in the 8 pathogen reactions were repeated or excluded. Pathogen detection was determined to be “positive” if the Ct value was below the cutoff Ct determined through limit of detection studies, described below, and if Ct confidence exceeded 0.15, and “negative” otherwise.

Pathogen-specific limit of detections were determined by spiking 5, 1.5 and 0.5 copies/µl of TrueMark Amplification Control I (1 x 10^5^ copies/μL) (Thermo Fisher Scientific) into pooled extracts from air samples collected for 4 days in an empty office room. The mean Ct value from the lowest dilution with detectable Ct in ≥75% of four replicate reactions was used as a cutoff for “positive” versus “negative” designation. In each case, this Ct was below the mean Ct detected in the no-template control.

### 2.3 SARS-CoV-2 Whole Genome Sequencing

SARS-CoV-2 multi-amplicon sequencing libraries were prepared from air sample nucleic acid extracts using the QIAseq DIRECT SARS-CoV-2 kit with optional Enhancer and Booster (QIAGEN) according to manufacturer instructions. Individual sample libraries were assessed for quality and quantity using a small-scale sequencing run on an Illumina iSeq, then re-balanced according to the number of SARS-CoV-2 mapping reads, and finally sent for deep sequencing on an Illumina NovaSeq X targeting 2 million clusters per sample (2×150 base reads).

WGS results were processed using the Freyja_FASTQ_PHB workflow available from Theiagen on the Terra platform [7, 35, 36]. Briefly, this workflow conducts read quality trimming and filtering using Trimmomatic and fastq-scan. Cleaned reads were then aligned with bwa to a SARS-CoV-2 reference genome (GenBank accession no. MN908947.3). Primer sequences were removed from the aligned bam file using iVar. Finally, Freyja was used to call demixed SARS-CoV-2 variants and determine their relative abundances [7]. Demixed results were downloaded locally and read into RStudio for processing and visualization [34]. Samples were included if their genome coverage was >50% and the negative controls (extraction blank) from that sequencing run had <50% coverage. Lineages were then aggregated into a select group of parent lineages for simplified visualization. Lineages that have been designated and tracked by the CDC COVID Variant Tracker (as of July 10, 2024) were selected as parent lineages, and aggregation assignments were determined using established Pango lineage designations [37]. Relative abundance plots were visualized using ggplot2. Merged air samplers were calculated by taking the average aggregated relative abundances of each lineage detected from the samples with data collected during that week.

### 2.4 Metagenomic sequencing with hybrid capture enrichment

Libraries were prepared for selected air samples collected from the hospital, storage room, and a water blank using the RNA Prep with Enrichment kit and Viral Surveillance Panel (VSP) (Illumina) according to the manufacturer’s instructions. In brief, 10 ng total nucleic acids from air sample extracts was subject to first and second strand cDNA synthesis and then tagmentated and amplified for 17 PCR cycles. Libraries were cleaned and normalized. VSP probes were hybridized to the libraries for 90 minutes then captured using magnetic beads. Enriched libraries were further PCR amplified for an additional 14 cycles and cleaned. Pre- and post-VSP capture libraries were sent for sequencing. Prepared libraries were sequenced on Illumina NovaSeq X, targeting 5-10 million clusters per sample (2×150 base reads) after library balancing, as described above.

Raw sequencing data was analyzed using the CZ-ID Illumina mNGS Pipeline v8.3 [38, 39]. In brief, raw data are uploaded to the CZ-ID web portal. Reads are quality and adapter trimmed using a customized version of fastp, human and duplicate reads are removed using Bowtie2 followed by HISAT2 alignment against reference human genomes and CZID-dedup, respectively. Reads passing quality and host filters are subsampled to 2 million paired end reads per sample. On average, 40% of data passing filters were included in the subsample. Reads are aligned to the NCBI NT and NR databases using Minimap2 and DIAMOND. Contigs are assembled using SPAdes and reads are mapped against contigs to assess coverage using Bowtie2. Contigs are aligned to a custom database of taxa identified through read mapping data using BLASTN and BLASTX. Identified taxa, number of reads and contigs aligning to taxa, percent identity to identified taxa, and E-values are summarized. Taxa were filtered to pathogenic human viruses as defined by CZ-ID. Taxa with fewer than 10 mapping reads were excluded. Low abundance taxa were manually verified using BLASTN of mapping reads. Total mapping reads and reads per million per sample/taxa were downloaded using the heatmap function within the CZ ID web interface.

Reference-based genome assemblies were prepared for adenovirus and influenza A using the TheiaCoV_Illumina_PE_PHB workflow available from Theiagen on the Terra platform [36, 40]. In brief, reads were adapter trimmed using BBDuk, quality trimmed using trimmomatic and host reads are removed using sra-human-scrubber. For influenza A, IRMA was used for genome assembly and characterization [41]. For adenovirus, reads were assembled to human mastenovirus B reference MW013769.1 using BWA and iVar. Genome coverage depth per site was generated using samtools. SARS-CoV-2 genome assemblies and lineage deconvolution was performed using Freyja, as described above.

### 2.5 Statistical analysis and comparison to clinical respiratory virus surveillance

For comparisons between air samples and clinical respiratory virus surveillance, United States clinical detection data were retrieved from the National Respiratory and Enteric Virus Surveillance System (NREVSS) [42]. Hospital-level clinical detection data were obtained from Rush University Medical Center Clinical Microbiology Laboratory. Statistical analyses were performed using GraphPad Prism (v. 10.2.2). Correlations were assessed using both simple linear regression and Spearman’s rank correlation coefficient with a two-tailed p-value (α = 0.05). Air detection data were smoothed using a second-order polynomial smoothing technique. Two-group comparisons were performed using Mann-Whitney test with a two-tailed p-value. Multiple group comparisons were performed using nonparametric Kruskal-Wallis test with Dunn’s correction and adjusted P values for multiple comparisons (α = 0.05).

## 3. Results

### 3.1 Respiratory pathogens are readily detected in hospital air samples

Air samples (n=318) were collected for analysis at 1-week intervals from samplers placed at 4 high-traffic hospital locations (Fig 1). Fourteen samples were excluded due to equipment failures during sampling. Forty weekly storage room samples were collected to assess environmental background in a room with very limited human activity. Weekly air samples were tested by qPCR for eight respiratory pathogens representing RNA and DNA viruses and a bacterium. 315 of 318 (99%) samples were positive for at least one respiratory pathogen, with a mean of 4.1 pathogens (95% CI: 3.9 – 4.2) detected in each specimen. Some pathogens were detected throughout the calendar year, with adenovirus, SARS-CoV-2, and *Streptococcus pneumoniae* detected in 83%, 97% and 96% of weekly air samples, respectively (Fig 2A). Others were seasonally detected, with human metapneumovirus, influenza A, influenza B, and respiratory syncytial virus (RSV) detected in 33%, 62%, 8%, and 38% of weekly air samples, respectively. Enterovirus was sporadically and rarely detected in 8% of weekly air samples. Air samples pathogen load was higher in samples collected from the ED compared to samples collected in hospital common areas, with weekly air sample Ct values significantly lower for 6 of 8 pathogens, all except influenza B and enterovirus, which had the fewest number of detections (Fig 2B). Mean Ct values of the positive control human RNaseP gene were not statistically different between ED and non-ED sites, indicating that higher pathogen load was likely due to the type of patients present in the ED and not higher human traffic near the samplers.

**Fig 2.**
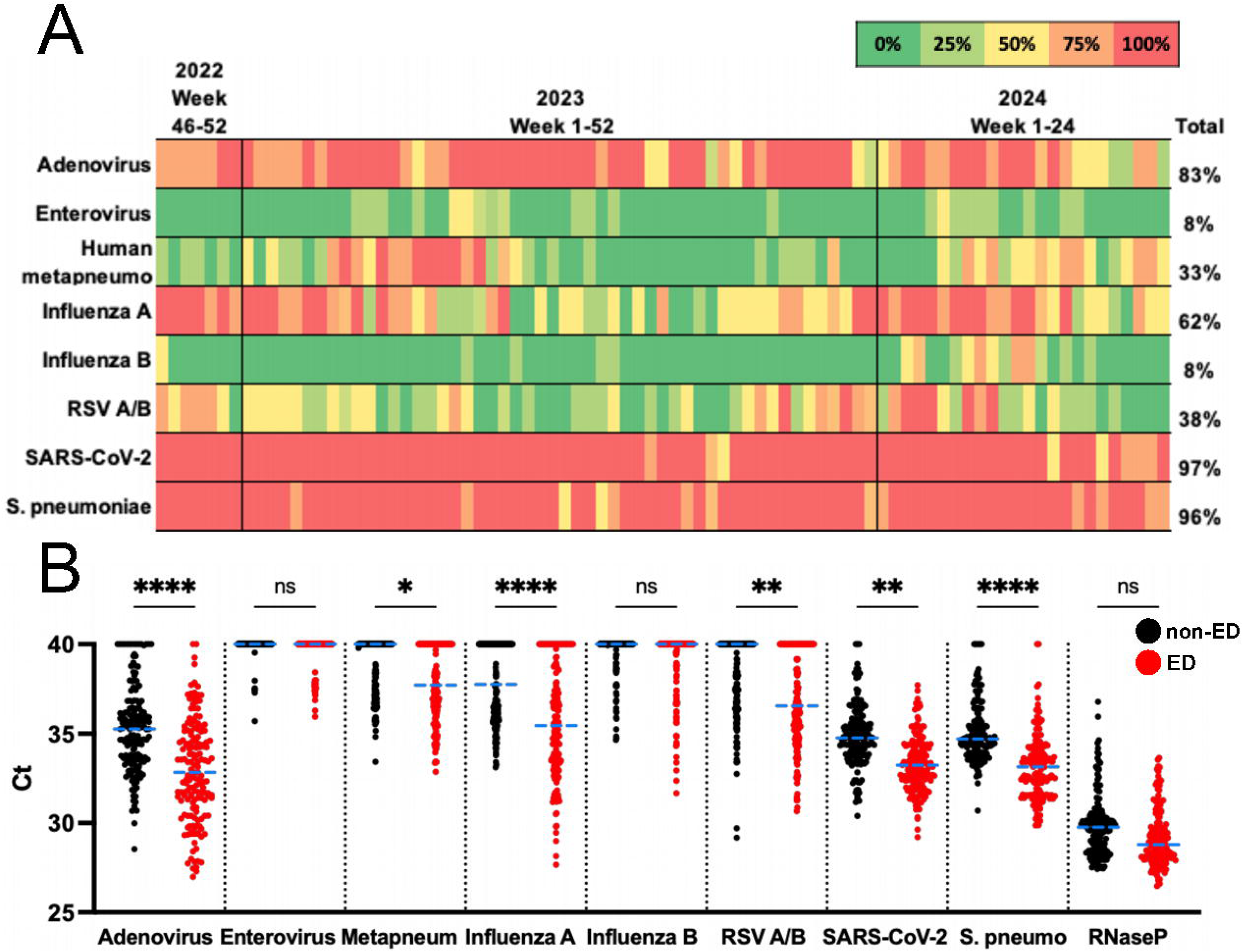
Respiratory pathogen detection in weekly hospital air samples. (A) The proportion of weekly samples (of four total) testing positive via qPCR for the indicated pathogens is shown by cell color. (B) qPCR Ct values for each weekly air sample from the emergency department (red) or non-emergency department (black) sites with mean indicated by blue dashed line. Statistical significance comparing ED and non-ED sites for individual pathogens is shown above; Kruskal-Wallis post-test (Dunn’s multiple comparisons test), * p<0.05, ** p<0.01, **** p<0001, ns = not significant.

### 3.2 Virus detection and load in air samples correlates with clinical respiratory sample positivity

To determine whether air sampling can mirror traditional clinical case-based surveillance, we compared rates of pathogen detection (air sample positivity) and pathogen load (air sample qPCR Ct values) to two sources of clinical surveillance data: i) clinical test percent positivity collected at the hospital where air sampling occurred and ii) percent positivity for US cases reported to CDC National Respiratory and Enteric Virus Surveillance System (NREVSS) [42]. We found that seasonal respiratory pathogens were strongly correlated between clinical and air surveillance. Peaks and valleys in local and national clinical percent positivity overlapped with air sample positivity trends for influenza A, influenza B, RSV, and human metapneumovirus (Fig 3A-D). Further, mean air sample Ct values dropped during periods of increased clinical percent positivity, indicating higher viral load in air samples collected during periods of higher community transmission (Fig 3A-D). Both air sample percent positivity and mean Ct value were strongly correlated to national and hospital percent positivity for these seasonal viruses, with Spearman r between 0.60 to 0.71 for influenza A, human metapneumovirus, and RSV air sample percent positivity and hospital/national percent positivity and −0.66 to −0.79 for air sample mean Ct values and hospital/national percent positivity (p<0.0001 for all comparisons) (Fig 3H-K, S1 Table).

**Fig 3.**
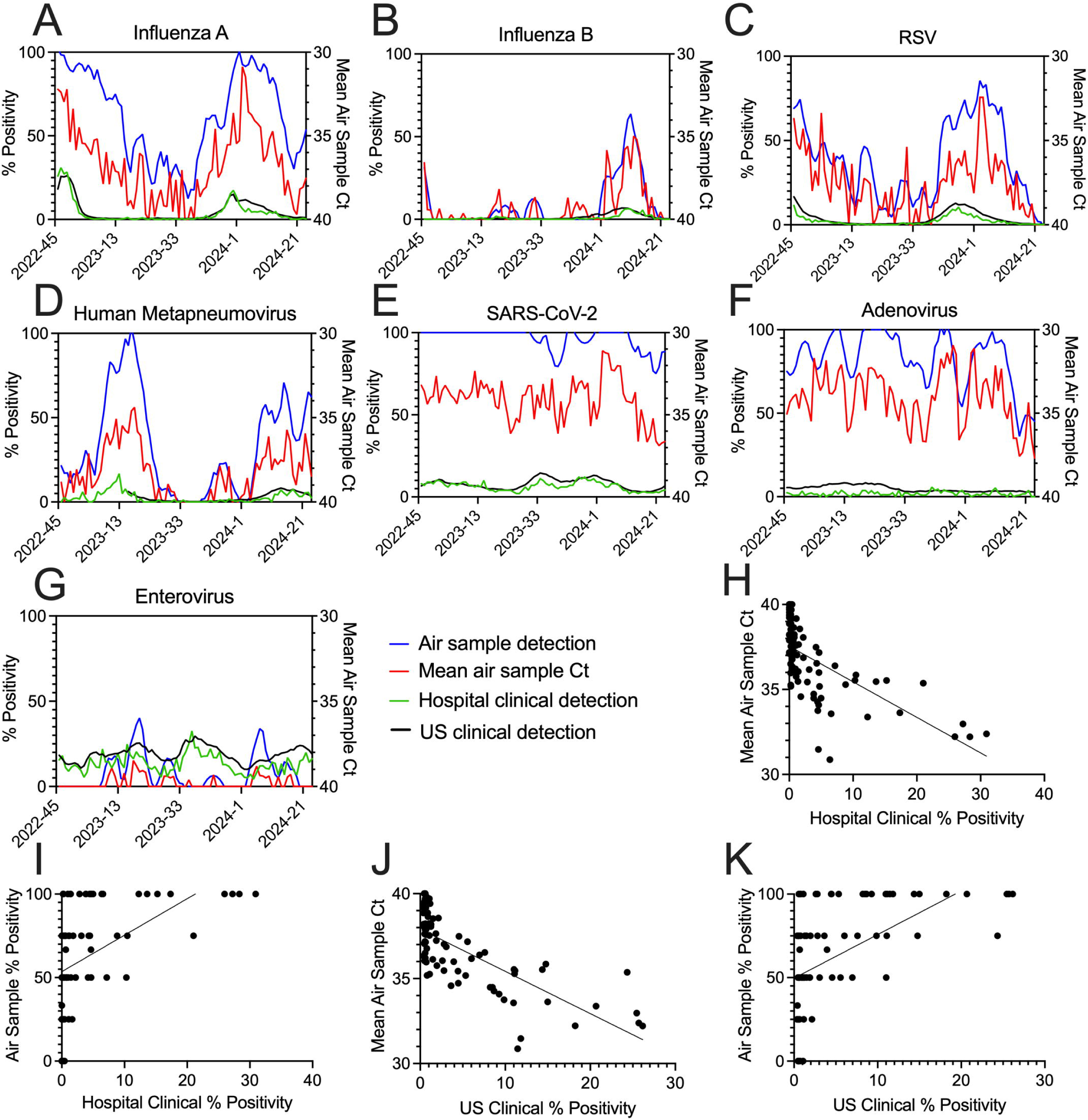
Virus detection and quantity in air samples correlates with results of clinical surveillance. (A-G) Weekly percent positivity for hospital clinical respiratory specimens (green, left axis) and US clinical respiratory specimens reported to CDC NREVSS (black, left axis) and air samples (blue, smoothed, left axis) is shown. For adenovirus, clinical percent positivity (hospital and US) is combined with rhinovirus. Mean of 4 individual air sample Ct values are also plotted for corresponding weeks (red, right axis, inversed). Sample collection week (x axis) is shown as collection year – CDC Morbidity and Mortality Weekly Report (MMWR) week. (H-K) Correlation between influenza A mean weekly Ct value and percent positivity and U.S. or hospital clinical percent positivity is shown with linear regression. Correlations for other pathogens are summarized in S1 Table.

SARS-CoV-2 was largely at saturation in weekly air samples with 97% positive during the study duration. However, a few periods of air sample detections falling below 100% overlapped with periods of declining national and local SARS-CoV-2 test positivity (Fig 3E). Indeed, there was a moderate correlation between SARS-CoV-2 mean air sample Ct value and hospital and national percent positivity (Spearman r=-0.44 and - 0.30, p <0.0001 and 0.0053, respectively, S1 Table). Adenovirus showed relatively minor changes in clinical percent positivity during the study period; air sample detections largely reflected this with weak but significant correlation between air sample mean Ct value and US clinical percent positivity and air detections and hospital/national clinical percent positivity (Fig 3F, Spearman r=-0.34 and 0.25 / 0.33, p=0.002, 0.024, 0.002, respectively, S1 Table). Enterovirus was the only virus without statistically significant correlation to clinical data. In this case, enterovirus clinical case positivity rates are combined with rhinovirus in both hospital and NREVSS surveillance, as clinical assays often do not distinguish between these two viruses. The assay used for air samples was enterovirus-specific. Contribution of rhinovirus to clinical data likely explains the lack of correlation to air enterovirus detections. Overall, this data suggests that hospital air sampling can effectively reproduce traditional clinical respiratory virus surveillance.

### 3.3 SARS-CoV-2 lineages in hospital air samples reflect regional clinical genomic surveillance

Genomic surveillance data can complement case surveillance by allowing characterization of pathogen variants and lineages, monitoring emerging lineages, and identifying mutations of interest. To determine whether SARS-CoV-2 genomes can be recovered from air samples, we subjected 204 air samples from all 4 sampling sites to SARS-CoV-2 tiled amplicon library preparation and sequencing. The bioinformatic tool Freyja, designed for SARS-CoV-2 wastewater sequence analysis was used to determine relative lineage proportions in air samples. We found that SARS-CoV-2 genomes could be readily recovered from air samples, with 178 (87%) yielding over 50% genome coverage and 117 (57%) yielding over 90% genome coverage. There was a strong correlation between percent genome coverage and air sample Ct value (Spearman r=-0.63, p<0.0001), with 95% of air samples exceeding our threshold for inclusion (>50% genome coverage) when Ct values were below 36 (Fig 4A). The mean SARS-CoV-2 Ct value was 34.1 for all air samples collected during the study period, with 87% below Ct 36, indicating that SARS-CoV-2 genomic surveillance would be successful for a large proportion of air samples collected in these settings (Fig 4B).

**Fig 4.**
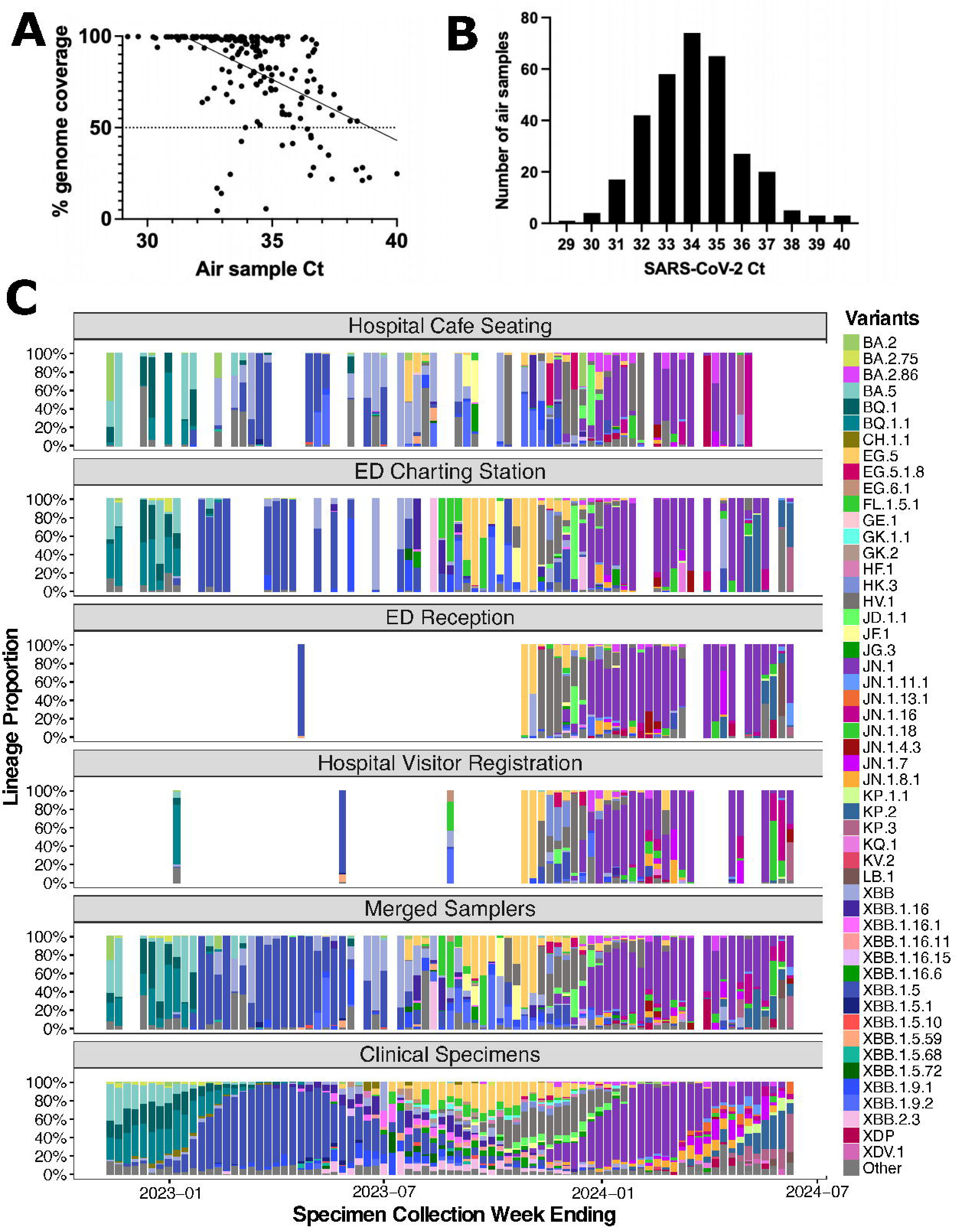
SARS-CoV-2 genomic surveillance using hospital air samples. (A) Correlation between percent SARS-CoV-2 genome recovered and air sample SARS-CoV-2 Ct value with linear regression. Dotted line indicates quality cutoff for inclusion in lineage analysis. (B) Histogram showing the number of air samples with varying SARS-CoV-2 Ct values. (C) SARS-CoV-2 lineage proportions detected in individual weekly air samples (top 4 panels), all available weekly air samples merged (5^th^ panel), and Chicago clinical specimens (6^th^ panel). Merged air samplers were produced by averaging detected lineage proportions from all available air samples that collection week. Clinical specimen lineage assignments were obtained from the Chicago Department of Public Health. Lineages reported are aggregated and colored according to the CDC COVID Data Tracker. “Other” designation indicates proportions defined as “Other” by bioinformatic tool Freyja as well as lineage proportions that are not able to be aggregated into CDC COVID Data Tracker-defined lineages (e.g. low abundance and/or recombinant lineages). Weekly proportions are shown, with the first MMWR week ending in January and July 2023 and 2024 indicated on the x axis.

Weekly air samples contained multiple SARS-CoV-2 lineages, with 99% containing more than one raw Pango lineage as reported by Freyja, and 95% containing more than one aggregated lineage following the CDC COVID Data Tracker lineage designations (Fig 4C) [37]. There were means of 20 raw and 6 aggregated SARS-CoV-2 lineages in air samples surpassing the genome coverage cutoff. To determine how lineages detected in hospital air samples compared to lineages detected by regional clinical genomic surveillance, we compared air sample lineage proportions to lineage proportions detected in genomic surveillance data collected by Chicago Department of Public Health in collaboration with the Regional Innovative Public Health Laboratory [43]. Lineages detected in air samples largely reflected contemporaneous lineages detected in regional clinical specimens, with lineage proportions merged from multiple air sampling sites most closely mirroring clinical lineage proportions (Fig 4C). Changing lineage proportion trends were clearly and simultaneously detected in air and clinical samples. For example, predominant BA.5 sublineages seen in late 2022 were replaced by XBB in the same weeks in early 2023 for air and clinical samples, and XBB sublineages declined while BA.2.86 and JN.1 lineages rose to predominance in overlapping weeks in late 2023 for both datasets. Notably, these data were achieved with 1-4 weekly air samples collected in a single hospital setting, whereas the clinical dataset contained mean 69 local specimens per week during this timeframe. Of 44 aggregated SARS-CoV-2 lineages emerging during the sampling period in clinical or air samples, the lineage was detected first in weekly clinical samples for 27 lineages (61%), 15 (34%) lineages were first detected in weekly air samples, and 2 (5%) were first detected in the same collection week for clinical and air samples. In sum, these data suggest that SARS-CoV-2 genomic surveillance can be effectively achieved from a small number of air samples collected in a hospital setting.

### 3.4 Many respiratory viruses are detected in air samples via metagenomic sequencing with viral enrichment

Metagenomic sequencing can allow simultaneous detection of many pathogens as well as detection of unexpected pathogens. However, recovery of virus genomes via metagenomic sequencing can be difficult from environmental and low biomass specimens. Viral enrichment can improve viral genome recovery from mixed samples. To determine whether metagenomic sequencing with viral enrichment could be used to detect and characterize viral pathogens from hospital air samples, we selected 24 air samples, including 11 collected in the ED, 9 collected from hospital congregate areas, and 4 storage room controls. Specimens collected during the 2022-2023 and 2023-2024 respiratory virus seasons (n=12) and off-season (n=12) were selected. Selected air samples were subjected to library preparation using hybrid-capture enrichment targeting 66 viruses of public health concern. We found that the percentage of sequence reads mapping to pathogenic human viruses increased from mean 0.001% pre-enrichment to mean 12.1% post-enrichment, representing a 10,000-fold increase in pathogenic virus-mapping data (Fig 5A). Respiratory viruses of interest were largely undetectable without enrichment, with 0 mapping reads per million for all 20 hospital air samples for adenovirus, influenza A, influenza B, human metapneumovirus, enterovirus and RSV and 19 of 20 samples for SARS-CoV-2. This increased to mean 5221, 1188, 209, 186, 14, 18 and 22 reads per million for SARS-CoV-2, adenovirus, RSV, influenza A, influenza B, human metapneumovirus and enterovirus respectively (Fig 5B). SARS-CoV-2 and adenovirus were detected in all 20 (100%) hospital air samples with enrichment, other respiratory pathogens were detected in 35% - 70% of air samples, with pathogen seasonality likely contributing to detection for some viruses.

**Fig 5.**
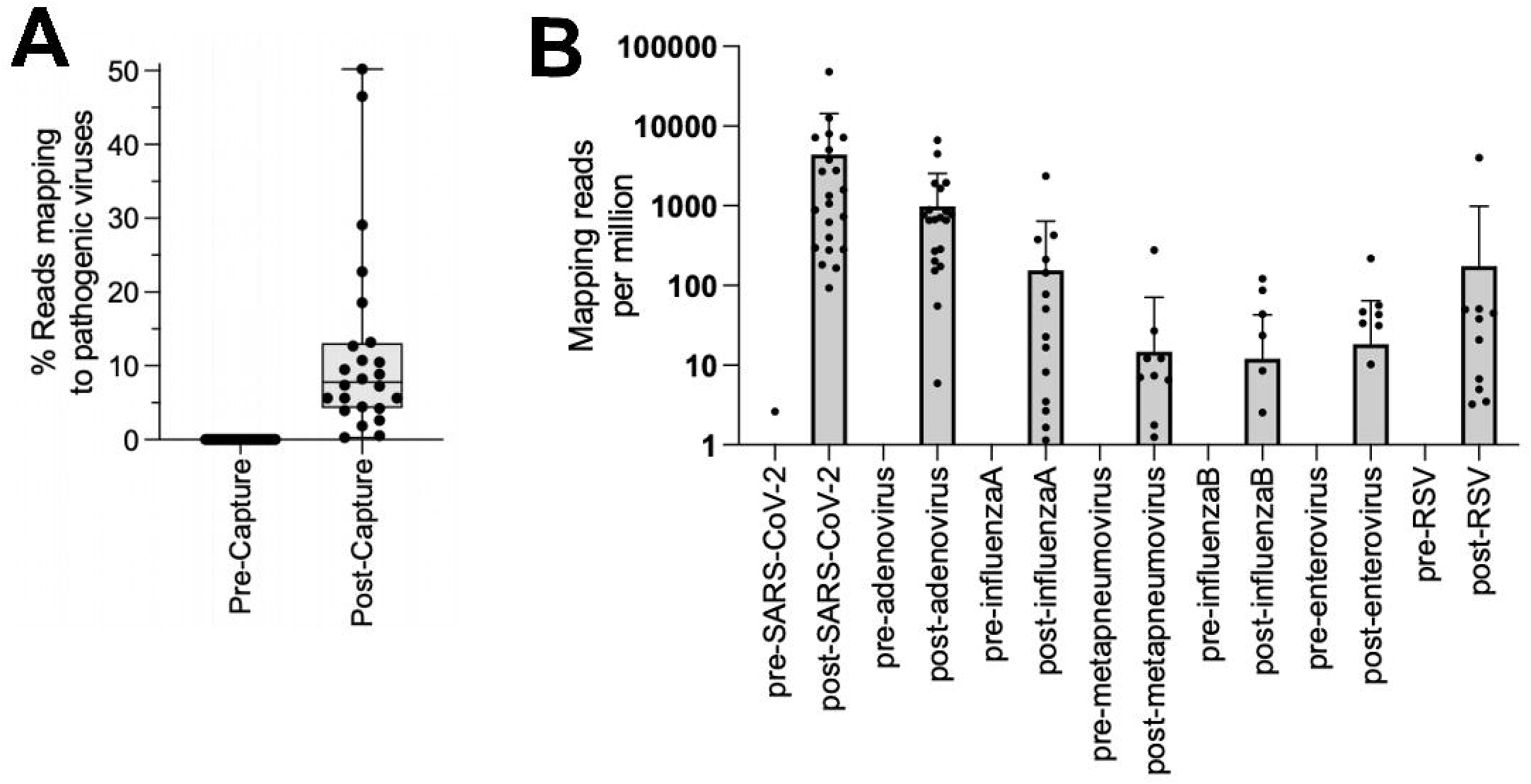
Enrichment of human viral pathogens in air sample metagenomic sequences. (A) The proportion of reads that mapped to human pathogenic viruses before and after viral hybrid capture enrichment is shown for n=24 air samples. (B) The number of reads mapping to specific viruses per million is shown for n=24 air samples before (“pre-“) and after (“post-“) hybrid capture enrichment. Samples with no mapping reads per million are not shown due to log scaling.

42 pathogenic human virus species were detected in hospital air samples with viral enrichment, including respiratory pathogens not targeted by qPCR, such as seasonal coronaviruses NL63, 229E, OC43, and HKU1, rhinovirus, parvovirus, parainfluenza virus and human respirovirus (Fig 6A). Several seasonal respiratory viruses were more commonly detected in air samples collected during respiratory virus season versus off-season. Pathogenic viruses typically associated with skin and enteric infections were also detected, including human polyomavirus 5 (Merkel cell polyomavirus), norovirus, mpox, and varicella zoster virus (Fig 6A). Pathogenic human viruses were detected frequently in the ED and hospital congregate areas, with mean 21.0 and 22.4 pathogenic human virus species detected in individual air samples from these sites, respectively. More pathogenic virus species were detected in air samples collected during respiratory virus season (mean 24.2) compared to during off-season in samples from all hospital sites (mean 19.3, p=0.002). Few pathogenic virus species were detected in air samples collected from a storage room (mean 3.0). Storage room specimens collected during respiratory virus season contained some respiratory viruses (e.g. coronaviruses), which may represent highly prevalent viruses at this time entering from the hallway, occasional staff access, or ventilation (Fig 6A). Storage room specimens during off-season contained only Merkel cell polyomavirus, which may represent ubiquitous presence or persistence within the built human environment. No viruses were detected in a water library blank included as a negative control. In sum, pathogenic human viruses can be readily detected in air samples via metagenomic sequencing with viral enrichment.

**Fig 6.**
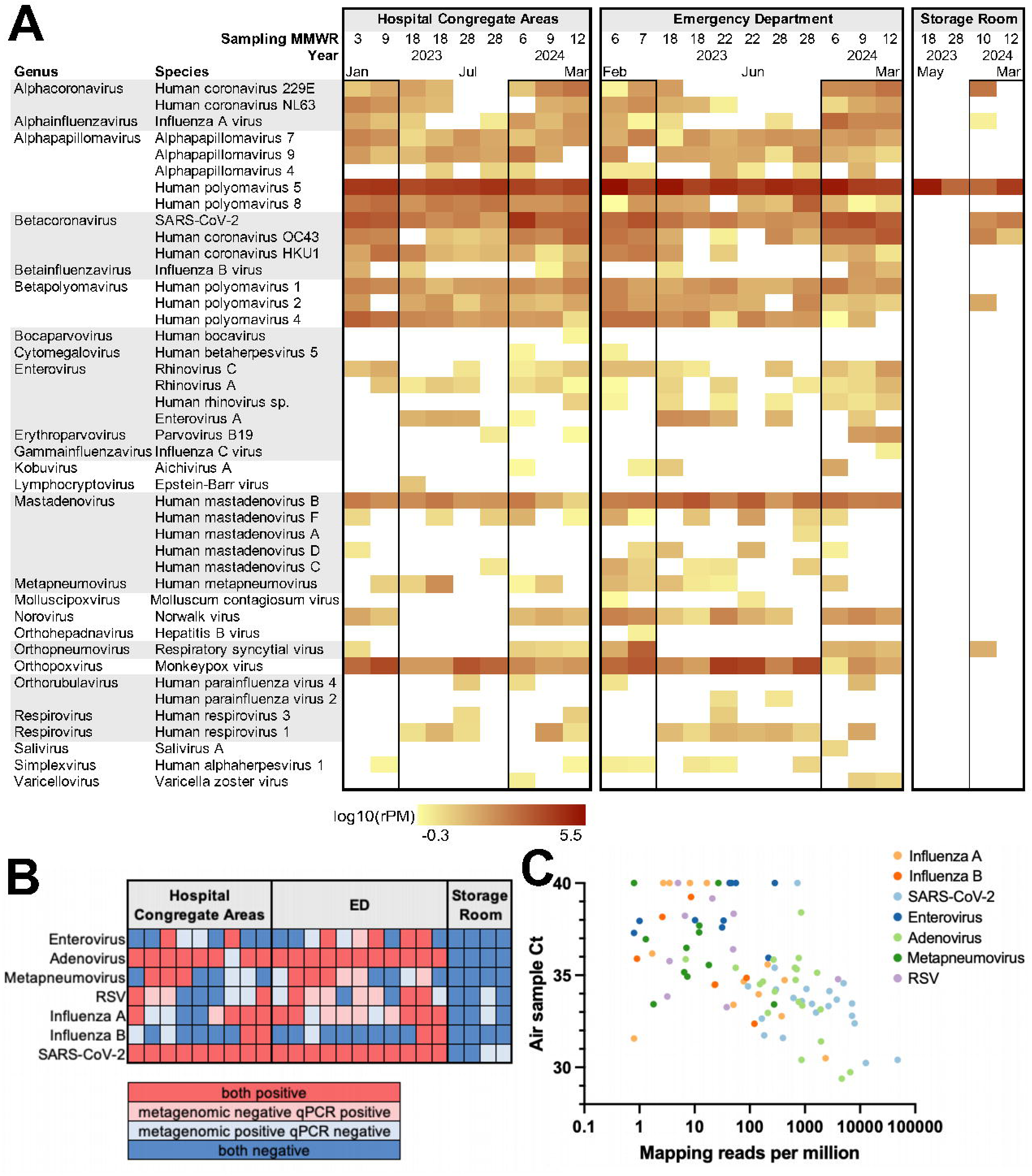
Many pathogenic human viruses are detected by metagenomic sequencing with enrichment. (A) Heatmap displaying the log10 reads per million mapping to pathogenic human viruses (rows) for individual air samples (columns). Air sampler location and collection year / MMWR week is shown above the columns; samples collected in the same week represent samples from two independent air samplers. Samples collected in respiratory virus season (October – March) are outlined with a box border. Respiratory-associated viruses are highlighted in grey. (B) Heatmap displaying concordance between viruses detected by both qPCR and metagenomic sequencing with enrichment. Air samples (columns) with sampling locations indicated at the top; red indicates the virus (rows) was detected by both qPCR and metagenomic sequencing (>10 mapping reads), dark blue indicates the virus was not detected by either qPCR or sequencing. Light red and light blue indicate discordant results, light red indicates the virus was detected by qPCR only, light blue indicates the virus was detected by sequencing only. (C) Association between air sample qPCR Ct value and mapping reads per million for the same pathogen is shown. Symbol color indicates the viral species.

Detection of selected respiratory viruses were largely concordant between qPCR and metagenomic sequencing with enrichment methods, with specimens positive or negative by both methods for 7 respiratory viruses described in Figures 1-2 in 77% of comparisons (Cohen’s kappa = 0.536, Fig 6B). In 23% of comparisons, detection of a specific virus differed between qPCR and metagenomic sequencing methods, with discordant positive detection more common with metagenomic sequencing (14%) than qPCR (9%). There was a strong correlation between air sample Ct value and mapping reads per million for the 7 respiratory pathogens quantified by qPCR (Spearman r= - 0.75, p<0.0001, Fig 6C). Air specimens with a Ct value below 35 had mean 2,681 mapping reads per million for that pathogen (range 0.01 – 48,164, 25^th^ – 75^th^ percentile 107.5 - 2,527). Overall, these data suggest that metagenomic sequencing with viral enrichment may allow for simultaneous surveillance of myriad human viral pathogens present in indoor air samples.

### 3.5 Complete viral genomes can be recovered from hospital air samples via metagenomic sequencing with enrichment

To determine whether virus genomes can be assembled from air samples via metagenomic sequencing with enrichment, we further analyzed three respiratory viruses: SARS-CoV-2, influenza A, and adenovirus. SARS-CoV-2 genome assemblies and lineage proportions were produced from metagenomic sequencing data. Of 20 air samples collected in the hospital, 13 (65%) had SARS-CoV-2 genome coverage above 50% and 3 (15%) had genome coverage above 90%, with mean 53.7% genome coverage (range 9.8 – 99.9%). Air samples collected in the storage room had the lowest SARS-CoV-2 coverage (range 0.1 – 1.0%). Of 13 samples exceeding 50% coverage via metagenomic sequencing, 11 had also been tested via SARS-CoV-2 amplicon sequencing with 10 exceeding 50% coverage for analysis inclusion. We found that within-sample SARS-CoV-2 lineage proportions detected by amplicon and metagenomic sequencing with enrichment in these 10 samples were highly concordant (Fig 7A). In one exception where a metagenomic specimen showed “Other” lineage designation compared to XBB and BA.5 sublineage designations via amplicon sequencing (ED specimen 6-2023, Fig 7A), the metagenomic specimen result was just above the 50% genome coverage inclusion cutoff.

**Fig 7.**
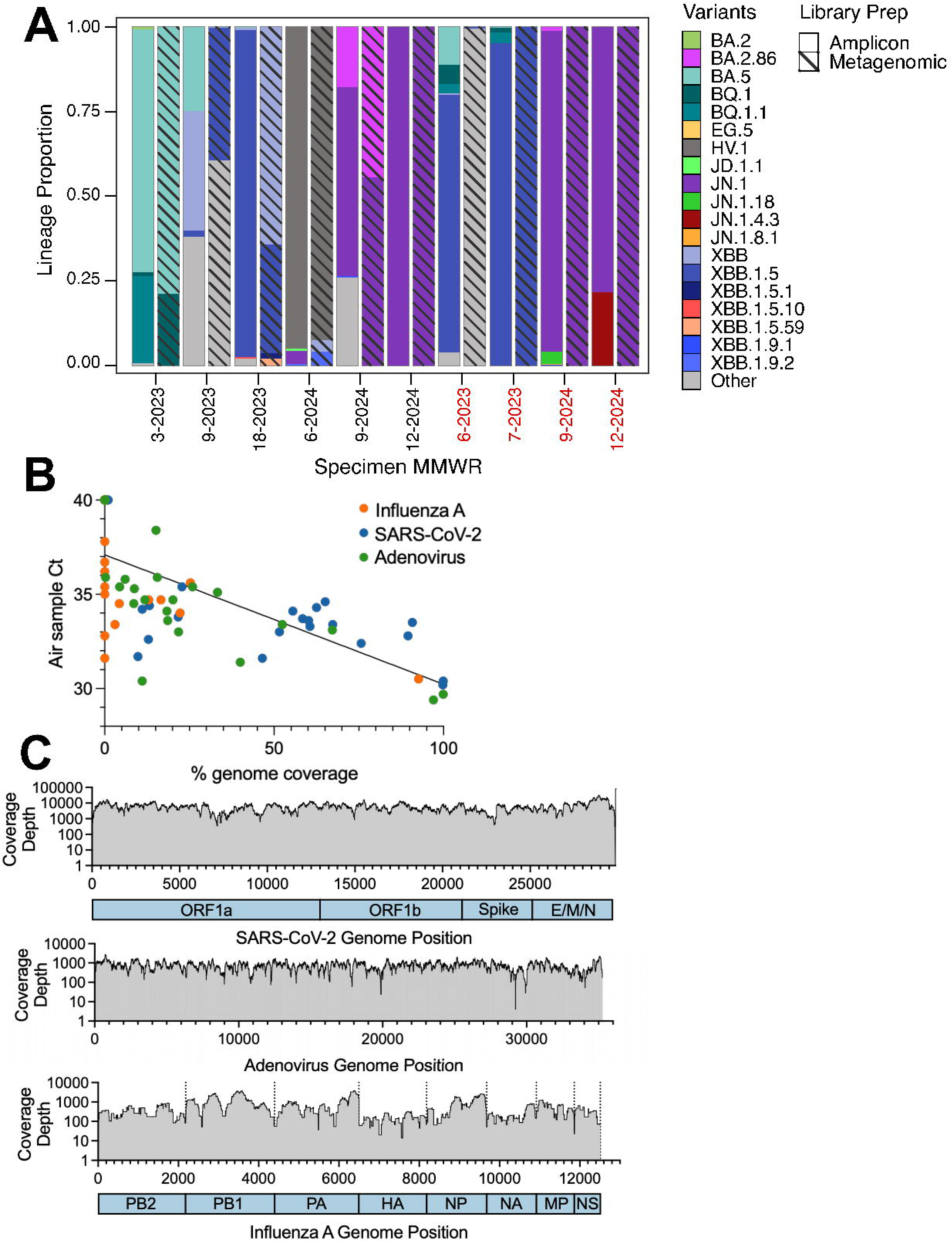
Respiratory virus genomes assembled from metagenomic sequencing data. (A) Relative SARS-CoV-2 lineage abundances detected in individual air samples collected in hospital congregate areas (black text label) or the ED (red text label). Specimen collection MMWR week and year are indicated. Lineage abundances for air samples prepared using tiled amplicon sequencing (solid bars) or metagenomic sequencing with viral enrichment (diagonal line patterned bars) are shown for air samples yielding >50% SARS-CoV-2 genome coverage by both methods. Pango lineages were aggregated according to those distinguished in the CDC COVID Data Tracker. (B) Association between air sample pathogen qPCR Ct value and percent genome coverage of reference-based assemblies produced from metagenomic sequencing with viral enrichment. Linear regression is shown, symbol color indicates viral pathogen. (C) Per nucleotide coverage depth for air samples yielding the highest-genome coverage assemblies for SARS-CoV-2 (top panel), adenovirus (middle panel) and influenza A (bottom panel). X axis indicates genomic position, horizontal bars below the x axis indicates genomic position of viral genes (SARS-CoV-2) or influenza A segments. For influenza A, vertical dashed lines indicate different genomic segments, which were concatenated for visualization.

Influenza A and adenovirus were also subjected to reference-based genome assembly. Air sample Ct value was strongly correlated with percent genome coverage (Spearman r = −0.74, p<0.0001, Fig 7B). Near-complete genomes were assembled for all three viruses from air specimens with Ct value ∼30 (Fig 7C). Mean coverage depths were 6,235 for SARS-CoV-2,761 for adenovirus, and 628 for influenza A for these specimens (Fig 7C). In all three assemblies, genomic sites with alternate variant frequency between 20 and 80% were identified, suggesting that multiple virus strains were sampled and sequenced. These data demonstrate that viral genomes can be assembled from air samples and used to characterize virus lineage and mutations of interest.

## 4. Discussion

This study supports air sampling as a viable pathogen surveillance tool. The data presented here suggest air sampling can be used i) as a complement or alternative to traditional clinical-based viral surveillance, ii) for efficient genomic surveillance of targeted pathogens using amplicon sequencing, iii) for pan-viral surveillance and/or genomic characterization using metagenomic sequencing with enrichment. Importantly, we find that pathogen positivity and load in air samples is well-correlated with national and institutional clinical percent positivity for multiple respiratory pathogens. Others have previously found that pathogens detected in air samples correlate with the presence of patients with known infections at the location [9, 14, 15, 44]. However, this is the first study to our knowledge to find that levels of multiple respiratory viruses of high clinical importance in air samples correlate with institutional and regional public health surveillance data. This critical data supports air sampling as a viable environmental surveillance method for infectious diseases.

Air sampling has the advantage over clinical testing-based surveillance in that it is not dependent on test-seeking behaviors or access to healthcare. Air sampling could thus reduce bias and improve equity in populations included in surveillance programs. While we targeted healthcare settings in this study, other highly trafficked sites may serve as good targets for surveillance of populations that are typically underrepresented in the hospital. Further, individuals shedding virus may contribute to air samples regardless of clinical symptoms. Indeed, we detected some seasonal pathogens like influenza A and RSV even when clinical percent positivity was low, indicating that air sampling may pick up asymptomatic cases or cases that are not tested for a specific pathogen given atypical timing. In addition, a single air sample can represent many patients, making air sampling more economical for viral surveillance than comprehensive clinical testing. Wastewater surveillance programs have been widely accepted in public health departments for these similar qualities. In comparison to wastewater, air sampling permits finer-scale population sampling, providing data at the building or room scale. Aggregate analysis of air samples from multiple buildings could enable neighborhood or city-wide surveillance with precisely defined boundaries, with the option of pooling air samples prior to testing for more cost-effective surveillance. Air sampling is also easier to coordinate than clinical or wastewater testing; an air sampler can be placed or moved within minutes, requires only an electrical outlet, and newer-generation air samplers require minimal training to operate, allowing staff of various backgrounds to execute sampling programs.

Here, air samples were collected in a large urban hospital and ED. Detection rates of respiratory pathogens were higher in this setting than other air sampling studies of community settings [9, 10, 45]. In our data, ED samples had the highest pathogen detections and quantities for most pathogens detected, posing this setting as an optimal one for air surveillance; sampling at a single ED site would have well reproduced the observed respiratory virus trends seen in combined sampling at four hospital sites. Hospital air surveillance data could be used as a reflection of community disease burden for public health or used internally by individual institutions to inform infection prevention practices (e.g. the need for masking during periods of high respiratory virus presence). Other sites that may benefit from air sampling include healthcare or long-term care settings with a low tolerance for infectious diseases, such as units housing immune-compromised individuals. In these settings, air sampling could be used as an early warning for the introduction of infectious diseases, though the frequency of sampling would need to be shorter than that employed here (e.g. daily) to be informative. We performed week-long sampling to support analysis of long-term trends. However, daily or sub-daily air sampling may be employed, with other studies showing detection of SARS-CoV-2 via air sampling in a matter of hours [8]. Indeed, sub-weekly sampling may be required for surveillance of SARS-CoV-2 rates in the hospital setting, as air samples in our study appeared saturated for SARS-CoV-2 in weekly samples. Air sampling could also be used for detection of rare or emerging pathogens at common points of entry (e.g. large international airports), or detection of spread of limited outbreaks to new populations. In these cases, comprehensive individual clinical testing for rare events may be difficult to coordinate or expensive to employ due to the large sample sizes needed.

The ability to sequence and assemble pathogen genomes from air samples further expands the utility of air sampling as a pathogen surveillance tool. Recovery of virus genomes from air samples using metagenomic sequencing has previously been considered prohibitively inefficient. Like others, we found that few viruses could be detected using metagenomic methods without viral enrichment, and those detected were at very low abundance, precluding assembly of informative genomes. However, viral enrichment using hybrid-capture of a panel of viruses was highly effective, allowing detection of 42 pathogenic viral species in air samples and recovery of many partial and full virus genomes. While metagenomic sequencing with target enrichment cannot be considered a truly ‘unbiased’ or ‘pathogen-agnostic’ method, new panels with hundreds to thousands of viral targets, including human and animal species, could offer near-pan viral enrichment [31]. Large panels of known pathogens may be sufficient to detect newly emerging and uncharacterized viruses, as even novel pathogens would presumably arise from evolution or recombination or zoonotic spillover of existing viruses. Viral enrichment using target-agnostic methods, such as clarification and/or filtration to remove large cells and nuclease treatment prior to nucleic acid extraction to remove non-encapsidated nucleic acids may further improve virus recovery from air samples but was not tested here [46]. In general, we observed better recovery of DNA over RNA viruses in metagenomic sequencing data, which is expected given the greater stability of DNA and more efficient recovery of DNA in sequencing library preparation, as it is not reliant on cDNA synthesis. Thus, methods to improve recovery of RNA viruses, such as sequence-independent single primer amplification methods may also be considered [29, 47].

Metagenomic sequencing of air samples could be used to characterize rare or emerging pathogens and aid in the selection or development of diagnostic assays for clinical testing. The ability to characterize virus lineages by metagenomic or targeted methods may also have utility in outbreak investigations, where the presence of single versus multiple genotypes could inform the heterogeneity of infections. Like another group, we also detected unexpected respiratory viruses like influenza C, a relatively understudied pathogen not subjected to routine surveillance or testing [29]. Detection of respiratory pathogens that may be excluded from clinical testing and/or surveillance underscores the utility of metagenomic sequencing for viral discovery and surveillance. Further, the proportion of metagenomic reads mapping to viral species was well correlated to virus levels determined by qPCR, which were in turn well correlated with clinical percent positivity data, suggesting that metagenomic sequencing could be used for pan-viral surveillance.

While many respiratory viruses were detected in hospital air samples, as expected, enteric and skin-associated viruses were also detected. Indeed, mpox was detected in 100% of hospital air samples subjected to enrichment sequencing and was one of the most abundantly detected viruses, but was not detected in storage room controls. Mpox has shown persistent transmission in Chicago since the 2022 global outbreak, including a local outbreak in 2023, with citywide cases detected in a majority of weeks sampled, thus its presence in this major Chicago-area hospital is not unexpected [48, 49]. Human skin is constantly shed into air and constitutes a large proportion of airborne dust particulates, explaining the ready detection of skin-associated viruses in air samples. Some skin-associated viruses such as mpox and varicella zoster virus can also cause systemic and/or respiratory infection, which may further explain their detection. Also, DNA viruses such as mpox are more likely to be stable in collected samples, and more readily detected by molecular methods employed here. Detection of enteric viruses may be due to fecal contamination of skin or presence of the virus in saliva for fecal-orally transmitted enteric viruses [50]. Others have described detection of enteric and skin-associated viruses in daycare, university and other congregate settings, suggesting that detection of these pathogens in air samples is not limited to healthcare settings [25, 29, 51]. Detection and genomic recovery of respiratory, skin, and enteric pathogens suggests that air sampling could be useful for surveillance and characterization of most pathogenic human viruses, not just respiratory viruses.

Caveats of our study include that this sampling method cannot distinguish between viable and non-viable viruses. Thus, detection of viruses in the air does not indicate transmission risk [52]. Also, the sampling method employed here likely inactivates viral particles and cannot be used to generate virus isolates for direct phenotypic testing and characterization. Specialized air samplers may be used to recover cultivatable viruses from the air but may be less efficient or accessible as a surveillance device [24, 53, 54]. Furthermore, qPCR Ct values were used as a proxy for viral load. Absolute quantification could be achieved using nucleic acid standards or digital PCR for more informative virus concentrations in air samples. Finally, quantification of pathogen load in air samples could benefit from normalization of relative human contribution to the samples, similar to human fecal normalization using foodborne viruses in wastewater studies. However, such normalization targets have yet to be developed for air samples.

Taken together, these data demonstrate that air sampling could be a paradigm-shifting tool for viral case and genomic surveillance, either as a complement to existing multifaceted surveillance programs, or as a more accessible and economical tool for smaller-scale environmental surveillance.

## Data availability

All raw sequencing data is available in the NCBI short read archive (SRA) database under BioProject PRJNA1148117. All other data is available at https://doi.org/10.5281/zenodo.14721822.

## Acknowledgements

This work was made possible by financial support through the Center for Emerging Infectious Diseases at Rush University Medical Center (1 GE1HS45832-01-00). We gratefully acknowledge Dr. Nicholas Moore and the Rush University Clinical Microbiology Laboratory for providing hospital respiratory virus clinical case positivity data. We thank Dr. Stephanie Black and the Chicago Department of Public Health Laboratory-Based Surveillance team and Regional Innovative Public Health Laboratory for SARS-CoV-2 lineage assignments from Chicago clinical specimens. We acknowledge Kevin Kunstman, Giancarlo Balangue, Felix Araujo-Perez, Jeremy Kahsen, Marisol Dominguez, and members of the Rush University Genomics and Microbiome Core Facility for air specimen collection and laboratory processing. We thank Chase Lodico and Dr. Galeta Carolyn Clayton for their support of this study. We gratefully thank Drs. David and Shelby O’Connor and Mitchell Ramuta from the University of Wisconsin Madison for sharing air sampling expertise and protocols in support of this study.

## Author contributions

H.J. Barbian, M. Gottlieb, S.J. Green, and M.K. Hayden conceived and designed the study and coordinated sampler placement. S. Bobrovska, E. Newcomer and H.J. Barbian collected and analyzed data. V.E. McSorely and A. Kittner coordinated and collected clinical SARS-CoV-2 genomic data. All authors were involved in drafting and revising the manuscript.

## Declaration of competing interest

The authors declare no competing interests.

---

**S1 Table:** Air sample and clinical surveillance correlation statistics.

| Pathogen | Correlation | Spearman r | P value |
| --- | --- | --- | --- |
| Adenovirus | Air sample Ct and hospital clinical % positivity | -0.2024 | 0.0664 |
|  | Air sample Ct and US clinical % positivity | -0.3351 | 0.002 |
|  | Air detection and hospital clinical % positivity | 0.2478 | 0.0239 |
|  | Air detection and US clinical % positivity | 0.3341 | 0.002 |
| Enterovirus | Air sample Ct and hospital clinical % positivity | 0.0237 | 0.8317 |
|  | Air sample Ct and US clinical % positivity | -0.0344 | 0.7578 |
|  | Air detection and hospital clinical % positivity | -0.047 | 0.6731 |
|  | Air detection and US clinical % positivity | 0.0093 | 0.9338 |
| Human Metapneumovirus | Air sample Ct and hospital clinical % positivity | -0.663 | <0.0001 |
|  | Air sample Ct and US clinical % positivity | -0.7872 | <0.0001 |
|  | Air detection and hospital clinical % positivity | 0.629 | <0.0001 |
|  | Air detection and US clinical % positivity | 0.709 | <0.0001 |
| Influenza A | Air sample Ct and hospital clinical % positivity | -0.759 | <0.0001 |
|  | Air sample Ct and US clinical % positivity | -0.7541 | <0.0001 |
|  | Air detection and hospital clinical % positivity | 0.6047 | <0.0001 |
|  | Air detection and US clinical % positivity | 0.6194 | <0.0001 |
| Influenza B | Air sample Ct and hospital clinical % positivity | -0.4822 | <0.0001 |
|  | Air sample Ct and US clinical % positivity | -0.5347 | <0.0001 |
|  | Air detection and hospital clinical % positivity | 0.4587 | <0.0001 |
|  | Air detection and US clinical % positivity | 0.4645 | <0.0001 |
| RSV | Air sample Ct and hospital clinical % positivity | -0.656 | <0.0001 |
|  | Air sample Ct and US clinical % positivity | -0.6805 | <0.0001 |
|  | Air detection and hospital clinical % positivity | 0.6233 | <0.0001 |
|  | Air detection and US clinical % positivity | 0.6256 | <0.0001 |
| SARS-CoV-2 | Air sample Ct and hospital clinical % positivity | -0.4362 | <0.0001 |
|  | Air sample Ct and US clinical % positivity | -0.3035 | 0.0053 |
|  | Air detection and hospital clinical % positivity | 0.1783 | 0.1069 |
|  | Air detection and US clinical % positivity | 0.069 | 0.5352 |

## Notes

### Competing Interest Statement

The authors have declared no competing interest.

